# The neural signature of methylphenidate-enhanced memory disruption in human drug addiction: a randomized clinical trial

**DOI:** 10.1101/2025.04.29.25326658

**Authors:** Ahmet O. Ceceli, Sarah G. King, K. Rachel Drury, Natalie McClain, John Gray, Priyanthi S. Dassanayake, Jeffrey H. Newcorn, Daniela Schiller, Nelly Alia-Klein, Rita Z. Goldstein

## Abstract

**Background:** Drug-related memories can hinder abstinence goals in drug addiction. Promoting non-drug memories via ventromedial prefrontal cortex- (vmPFC) and amygdala-guided extinction yields mixed success. Post-retrieval extinction (RE) destabilizes and updates memories during reconsolidation, improving extinction. Supplementing RE, we tested methylphenidate (MPH), a dopamine-agonist that promotes PFC-dependent learning and memory in cocaine use disorder (CUD). In an Early Phase 1 double-blind randomized clinical trial using a within-subjects design, participants received oral MPH (20 mg) or placebo before the retrieval of some of the conditioned stimuli (i.e., reminded CS+ vs. non-reminded CS+) followed by extinction; lab-simulated drug-seeking was measured the following day.

**Results:** Lower vmPFC activity following non-reminded CS+ (standard extinction) under placebo replicated the putative impairments in CUD; separately, RE (trend) and MPH conditions recruited the vmPFC, and RE’s vmPFC-reliance correlated with drug-seeking only under placebo. Crucially, MPH-combined RE normalized cortico-limbic processing, bypassing the vmPFC and its amygdala connectivity.

**Conclusions:** Pharmacologically-enhanced drug memory modulation may inform intervention development for addiction recovery.

## Introduction

Cocaine addiction is a persistent and cyclical brain disease with no FDA-approved pharmacotherapies for its treatment^1^. Cues that have been previously associated with drug stimuli and use, such as drug paraphernalia or locations, can acquire incentive properties as conditioned stimuli (CS)^2^. Via mesocorticolimbic dopaminergic circuitry including the ventromedial prefrontal cortex (vmPFC), amygdala, nucleus accumbens and ventral tegmental area, these Pavlovian (conditioned/automatic) processes may precipitate intense physiological states such as craving and promote drug seeking and relapse despite long periods of abstinence^3^. Therefore, modulating these drug related memories has been a central target for enhancing drug addiction recovery^4^.

Cue exposure-based therapies, rooted in vmPFC-mediated Pavlovian extinction, aim to reduce maladaptive conditioned responses. Typically in extinction studies, the CS is presented in the absence of the unconditioned stimulus (US) to which it was previously paired, forming a new CS-no US association as accompanied by an increase in vmPFC signaling^5^. However, extinction does not erase the original CS-US memory, rendering it susceptible to reemergence via spontaneous recovery due to passage of time or reinstatement upon reencountering the US (see ^6^ for a review). In addition, both the integrity and function of the vmPFC are compromised in individuals with drug addiction^7–10^. For example, we reported lower vmPFC gray matter volume^11^ and decreased vmPFC activity during standard drug-cue extinction and its next-day retention^12^ in individuals with cocaine use disorder (iCUD) as compared to healthy controls (HC). Similarly, lower vmPFC function has been reported during extinction of aversive shock-associated cues in iCUD as associated with higher physiological arousal to the CS following extinction^13^. Given these vmPFC deficits and the limited success of extinction-based therapies in preventing relapse in drug addiction^14^, a need for alternative neuroscience-informed strategies, especially those that normalize and/or bypass vmPFC function, is evident.

Leveraging a reconsolidation window during which memories are labile for alteration, a candidate approach is post-retrieval extinction (RE). Here, a CS is presented (i.e., “reminded”) after its association with the US has been encoded, destabilizing the original CS-US memory prior to extinction learning (i.e., CS-no US)^15–19^. In the general population, RE outperforms standard extinction in the reduction of conditioned responses to fear-associated CS, with lower spontaneous recovery of skin conductance responses to the reminded compared to the non-reminded CS the next day^20^. Compared to standard extinction, vmPFC activity and its connectivity with the amygdala decreases during RE, suggesting that this behavioral manipulation is potentially suitable for populations with compromised vmPFC function^21^ and structure^11^. In preclinical studies, RE is associated with decreases in conditioned responses during cocaine, methamphetamine, and heroin cue reinstatement, cocaine and methamphetamine spontaneous recovery and in subsequent drug seeking^22–24^. Extending these patterns to humans, individuals with nicotine or heroin addiction who underwent RE reported lower cigarette or heroin cue induced craving—effects that were evident up to one or six months later, respectively^22,25^. However, the role of the vmPFC (and its connectivity with the amygdala) in this putative RE modulation of salient drug memories in human drug addiction is yet to be studied. Its pharmacological modulation also remains to be explored, of importance for the future design of optimized interventions.

Pharmacological interventions to alter drug memories during reconsolidation, such as with N-methyl-D-aspartate (NMDA) or beta-adrenergic receptor based agents (including the agonist propranolol), show promise in animal studies^26–30^, as well as in humans yet with mixed results^22,31,32^. For example, memantine (an NMDA receptor antagonist) administered to cigarette smokers prior to a cigarette memory reactivation procedure did not improve physiological, craving, or relapse latency measures compared to placebo^31^. In contrast, smokers who received propranolol (40 mg oral) 1 hour before the retrieval of a nicotine US reduced subsequent cue-induced craving as compared to a control condition (a delayed propranolol dose given 6 hours after retrieval)^33^. In a similar vein, people with heroin addiction who received propranolol (40 mg oral) a day after learning a word list comprising heroin and non-heroin related words, remembered fewer drug words (but comparable neutral words) the following day—an effect dependent on word reactivation (writing down the learned words)^32^. However, given the psychotomimetic side effects (especially of NMDA-based agents and, less frequently, beta-adrenergic agents in higher doses), the translational value of these results is limited^34–36^.

Commonly used to treat attention-deficit/hyperactivity disorder, methylphenidate (MPH) may serve as a feasible candidate to enhance RE in drug addiction. Similarly to cocaine, and as a partial agonist, MPH binds to dopamine transporters, increasing extracellular dopamine (and norepinephrine) concentration in the brain^37^. However, its rate of clearance when orally administered is slower, not inducing craving or a high, thereby associated with low abuse potential and viability as a therapeutic agent^38,39^. In rodents, MPH facilitates learning-induced cortico-limbic plasticity^40^, memory consolidation^41,42^, and extinction learning via dopamine receptors^43^. In humans, a low oral dose of MPH reduced impulsivity and normalized vmPFC function during a drug-cue reactivity task in iCUD^44^; it also enhanced its resting-state functional connectivity with the striatum, reducing abnormal hyper-connectivity within striatal subregions as associated with severity of addiction^45^. Thus, potentially via dopaminergically-mediated normalizations in neural processing, encompassing task-relevant enhancements^38^, MPH may also optimize activity in regions that support the decoupling between the CS and US during RE.

To our knowledge, no study to date has inspected the neurobiology underlying a MPH-enhanced RE modulation of drug memories in substance use disorders either in humans or using preclinical models of drug addiction. Using a novel fMRI task that captured brain activity during RE (and standard extinction), in a placebo-controlled, double-blind, within-subject, randomized clinical trial (ClinicalTrials.gov ID: NCT05978167) we administered MPH (20 mg oral) timed to reach peak effects during the retrieval of a drug-cue memory in iCUD. As visualized in Figure 1, we hypothesized each separate component in this combined pharmacological-behavioral approach to modestly change vmPFC function, exemplified with increases over the course of extinction, mirroring patterns during standard extinction in the general population^5,21^. Crucially, we hypothesized a normalizing effect on vmPFC function (and connectivity with the amygdala and other mesolimbic regions) for the combined MPH-enhanced RE effect, yielding clinical utility as reflected in brain-behavior associations with next-day drug seeking.

**Figure 1.**
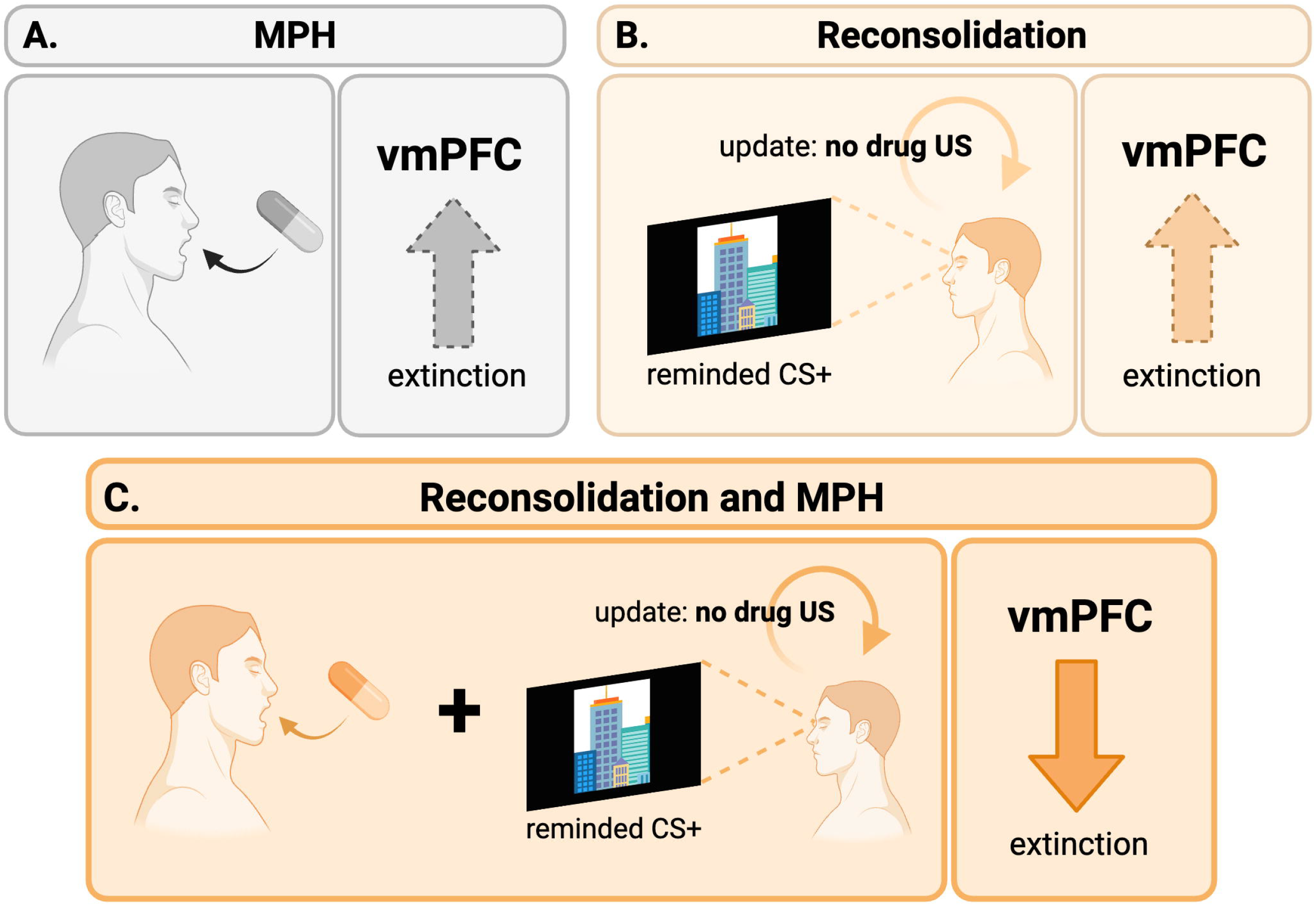
A schematic representation of the separate effects of methylphenidate (MPH; panel A), reconsolidation (panel B) and their combination (panel C) on ventromedial prefrontal cortex (vmPFC) function during extinction in individuals with cocaine use disorder. Separately, we expect MPH and reconsolidation manipulations (depicted in gray and orange, respectively, as consistent with non-reminded and reminded CS+ results in Figure 3) to induce modest vmPFC increases during extinction (as indicated by the dashed arrow and lighter shade). When combined, we expect a disengagement of the vmPFC, in line with patterns observed in the general population. US: unconditioned stimulus.

## Methods

The Icahn School of Medicine at Mount Sinai’s Institutional Review Board approved all study procedures. All participants provided written informed consent prior to participating in the study and were compensated after study completion. Recruitment spanned July 10, 2023 to July 29, 2024, ending upon reaching the target sample size.

### Participants

A power analysis of our previous publication indicating RE-related vmPFC effects in the general population ^21^ yielded an effect size of d=0.42; our previous results revealing MPH vs. placebo mediated vmPFC changes in a CUD sample, undergoing a comparable fMRI drug cue task^44^, yielded an effect size of d=1.4, justifying the sample size. Twenty iCUD were recruited via inpatient and outpatient addiction treatment facilities, flyers, and word of mouth. Two individuals’ data were omitted due to incidental MRI findings, and thus the analyses include the remaining 18 participants (mean age: 40.6±8.7, 6 women). A comprehensive clinical diagnostic battery (inclusive of the MINI International Neuropsychiatric Interview^46^ and the Addiction Severity Index^47^), medical screen, and electrocardiogram, conducted by trained research staff under a physician’s and a psychologist’s supervision, determined eligibility and drug use severity (see Supplementary Materials for details and exclusion criteria). All participants met DSM-5 criteria for CUD with cocaine as the primary drug of choice, with five individuals also presenting a current opioid use disorder diagnosis; all other psychiatric comorbidities including other substance use disorders that are commonly observed in people with drug addiction (e.g., to opioids, alcohol, tobacco, or cannabis) were either in partial or sustained remission. Table 1 details demographics, cocaine addiction severity measures in addition to opiate, alcohol, tobacco and cannabis use. Fifteen participants underwent medication-assisted treatment [14 methadone (mean dosage at baseline 110.71±42.92 mg), 1 buprenorphine/naloxone (16 mg)]; with the exception of one participant (increases from 120 to 140 mg), methadone dosages between pill administration days were identical.

**Table 1.** Sample profile.

| **Variable** | **Mean** | **Std. Dev.<sup>1</sup>** |
| --- | --- | --- |
| Age | 40.6 | 8.7 |
| Sex (Male/Female) | 12/6 |  |
| Race (Black/White/Other) | 5/11/2 |  |
| Education (Years) | 12.2 | 1.6 |
| Verbal IQ (WRAT-IV, Scaled) | 99.6 | 15.4 |
| Non-Verbal IQ (Matrix Reasoning, Scaled) | 10.2 | 3.1 |
| Beck's Depression Inventory II | 11.1 | 11.4 |
| Beck's Anxiety Inventory | 10.3 | 11.1 |
| Severity of Dependence Scale | 8.8 | 4.2 |
| Cocaine Selective Severity Assessment | 16.1 | 17.1 |
| Cocaine Craving Questionnaire | 10.3 | 13.2 |
| Cocaine Regular Use (Years) | 9.5 | 8.1 |
| Cocaine Heaviest Use (Years) | 4 | 4.3 |
| Heroin Regular Use (Years)<sup>2</sup> | 9.3 | 6.9 |
| Heroin Heaviest Use (Years)<sup>2</sup> | 3.8 | 2.2 |
| Cigarette Smoking (Current/Past/Never) | 16/1/1 |  |
| Fagerström Test for Nicotine Dependence | 3.3 | 1.9 |
| Alcohol Use to Intoxication (Years) | 6.5 | 9.5 |
| Marijuana Regular Use (Years) | 9.3 | 10.4 |

<sup>1</sup> Std. Dev: standard deviation.
<sup>2</sup> Data for heroin-related variables did not include two participants due to lack of heroin use history.

#### Learning and Neuroimaging Procedures under MPH or Placebo

In each of the two fMRI sessions, participants were randomized (allocations performed and concealed by independent staff) to orally receive MPH (20 mg) or placebo (lactose) in identically looking capsules. About four hours prior to pill administration, all participants underwent a Pavlovian conditioning task (outside of the MRI), as depicted in Figure 2. Here, two sets of distinct neutral visual cues (conditioned stimuli: CS+ and CS-) were partially (38%) reinforced with a clip of drug use (i.e., smoking a crack cocaine pipe) or a matched non-drug action (i.e., holding a pen near mouth) US, respectively. The non-reinforced trials only presented the CS in absence of the US. The CS were cartoon images of groups of people or buildings; analyses here collapse across these social/non-social cue types (their comparison will be reported elsewhere). In each trial, the CS remained on the screen for 4 seconds, and in reinforced trials, co-terminated with the 3-second US. The trial ended with an inter-trial interval that randomly varied between 5 and 7 seconds. We used six distinct CS images to encompass the social/nonsocial and reminded CS+/non-reminded CS+/CS- (see below) factors, presented in a pseudorandomized order (no more than two consecutive trials with the same image). This pseudorandomization was not applied to 7 participants due a script error. The conditioning task was delivered over three identical runs, each containing 48 trials (16 reminded CS+, non-reminded CS+, and CS-), for approximately eight minutes per run. A classic Flanker task was integrated into the conditioning task to increase task engagement (and thus not analyzed). Here participants were instructed to select the direction of the middle arrow among the five arrows that occasionally appeared on the screen as quickly and accurately as possible using the corresponding key on the keyboard. These Flanker trials were randomly placed at the end of 25% of the task trials with a 1-second response window. The task was coded using E-Prime (v3, *Psychology Software* Tools).

**Figure 2.**
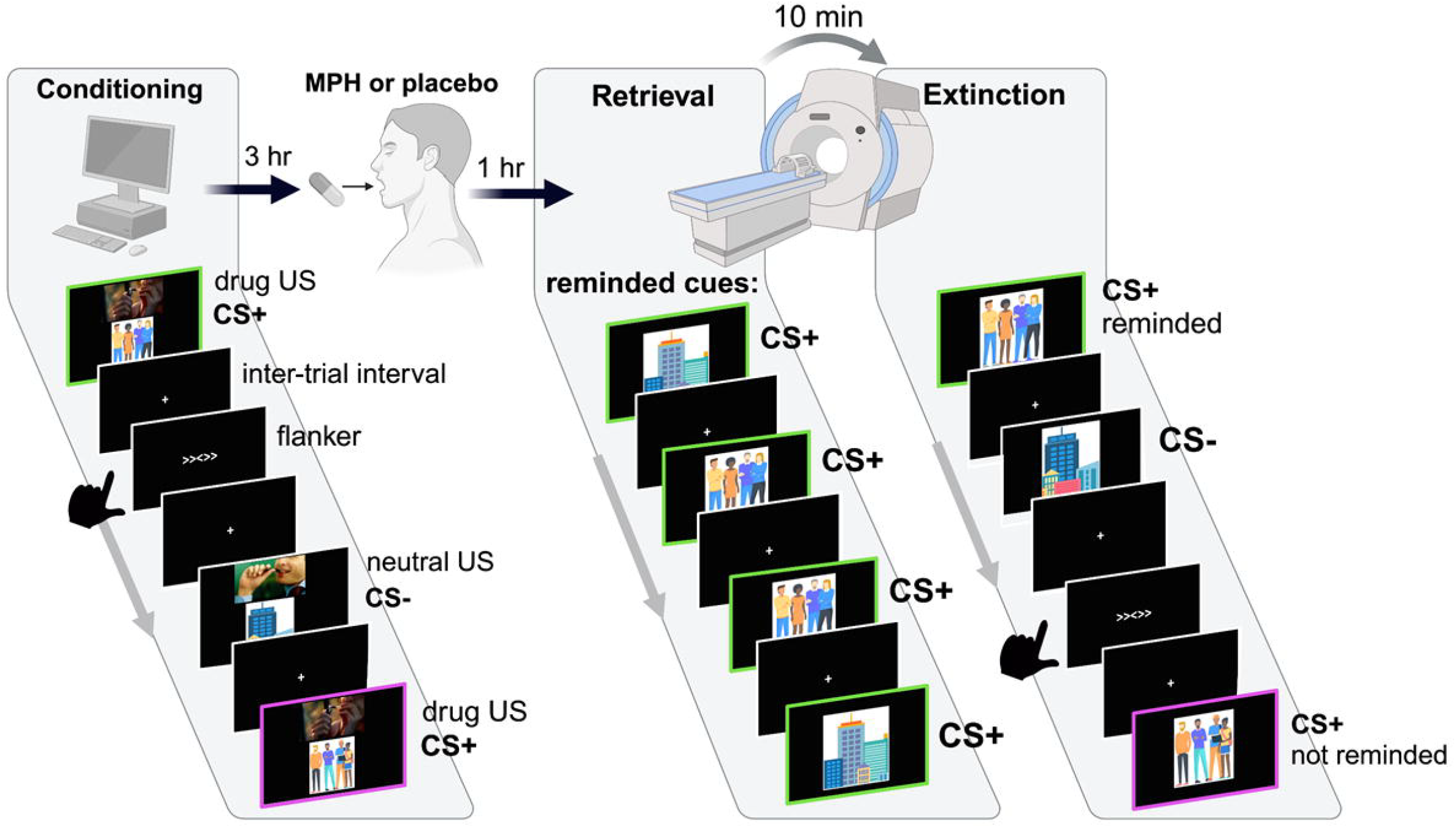
Study design. Participants underwent Pavlovian conditioning with pictorial conditioned stimuli (CS) and a brief video clip of drug use as the unconditioned stimulus (US); CS-was paired with a matched nondrug clip. Flanker trials served to improve engagement with the task. Three hours later, participants received either 20 mg methylphenidate (MPH) or placebo. One hour later, some of the conditioned stimuli were presented (while undergoing MRI) to induce retrieval. Ten minutes later, an extinction task comprising the reminded and non-reminded CS+ (as well as the CS- and flanker trials) followed. The next day, participants underwent an identical extinction task (i.e., “re-exposure”) outside of the MRI and completed a picture choice task that estimated lab-simulated drug-seeking (not pictured). These procedures were repeated approximately one-week later with the remaining pill.

Following prior procedures^12^, immediately before and after the conditioning and after the next-day extinction retention task (see below), participants provided subjective CS and US ratings using a 1-9 scale for valence (pleasantness) and arousal, and craving intensity for the drug US. Participants also completed a two-alternative forced choice task whereby they chose for preference of viewing between the two CS or US images in all possible pairs (e.g., a drug-associated CS or a neutral-associated CS). The drug US was rated to have lower valence and higher arousal than the neutral US as expected^48,49^, and the non-reminded CS+ was rated to have lower valence than the CS- (reminded CS+ being comparable to both), attesting to a potent US and the success of conditioning. Importantly, ratings did not yield significant differences between pill conditions in CS or US related valence, arousal, or craving intensity (for drug US) before or after the fMRI task. A trend for a lower craving in response to the drug US under MPH compared to placebo was non-significant, *F*(1,79)=3.86, *p*=.053, and not specific to a learning phase (e.g., before or after extinction). The means and standard deviations of these ratings are detailed in the supplement (all ps>.108). Results of the choice preferences also were unremarkable (see supplement).

An hour after pill administration (i.e., during peak MPH concentration^50^) and during fMRI, participants were presented two trials of the CS+ that was designated as the reminded CS+, but not the other CS (non-reminded CS+ or CS-), to induce reconsolidation (i.e., retrieving the reminded CS+ memory and updating it with a no-US association). Ten minutes later (as consistent with reconsolidation practices^20,21^), participants underwent extinction over the course of three runs. Task parameters and length of the extinction task were identical to the conditioning task described above, except none of the trials co-terminated with a US and subjective ratings/choice were not collected before or after the task. After the MRI scan (within 90 minutes of pill administration), 2 mL of blood was drawn for estimating blood MPH concentration. Participants were asked to guess whether they took the MPH or placebo pill at the end of the study day. The above procedures were repeated in their entirety in an identical study session approximately a week later (mean days between sessions: 8.8±4.5), at the same time of day, to study the effects of the remaining pill. Supplementary Table 1 details affective (collected before pill administration), physiological (cardiovascular measures collected 3 hours before and the day after pill administration as part of daily medical screening), and drug use recency measures (as well as guessing accuracy for pill taken) for the two pill administration days, none of which showed significant differences between days (all ps>.332). See Supplementary Table 2 for urine toxicology results for these study days.

#### Next-Day Drug-Biased Behavior Assessments

Participants returned the next day for estimates of drug-biased behavior. First, all participants underwent an extinction procedure identical to the previous day’s task except it was performed outside of the MRI scanner. This re-extinction step served to minimize potential effects of spontaneous recovery on the behavioral drug seeking measure of interest (see below). Subjective valence and arousal ratings and forced-choice were collected only after re-extinction to avoid reinstatement. Next, we estimated drug-seeking (a drug biased behavior) via a well-validated picture choice task where we sampled behavioral choice to view pleasant (e.g., people smiling), unpleasant (e.g., wounds), neutral (e.g., office supplies), and drug picture stimuli (e.g., people simulating the use or preparation of crack-cocaine)^51–53^. Consistent with the impaired response inhibition and salience attribution model (which emphasizes the excessive salience attributed to drug cues at the expense of alternative reinforcers in drug addiction)^7^, and given its concurrent and prospective prediction of cocaine use in iCUD^52^, drug vs. pleasant picture choice frequency served as an index of simulated drug seeking.

### Data Analyses

#### Blood MPH Concentration Estimates

Blood plasma was separated from whole blood within approximately 15 minutes from sample collection and stored at -80°C until analysis. MPH was extracted from plasma and the extracts were derivatized before gas chromatography/tandem mass spectrometry. The isotope dilution internal standard calibration method was used to accurately calculate MPH estimates^54^ (see Supplementary Materials for details).

#### Magnetic Resonance Imaging

A 3T Siemens Skyra was used for structural and functional imaging with the following time to repetition and time to echo parameters: 2,400/2.07□ms and 1,000/35□ms, respectively (see the Supplementary Materials for further acquisition and data preprocessing details). The MPH and placebo scans were comparable in mean framewise displacement [MPH: 0.34±0.21 mm, placebo: 0.42±0.36 mm, *t*(27.17)=0.82, *p*=.417).

#### BOLD-fMRI Data Analyses

All CS events (i.e., reminded CS+, non-reminded CS+, CS-) were sampled from their onset to offset (4 s) and convolved with a double-gamma hemodynamic response function to be included in the general linear model (GLM) along with their temporal derivatives using FSL’s FEAT (version 6.0.7.4^55^). Inter-trial intervals and Flanker events contributed to the implicit baseline.

The fMRIprep confound timeseries (see supplement) were included as nuisance regressors. The reminded>non-reminded CS+ contrast map served as the index of RE-related brain activity. These first-level maps were entered into fixed effects models to generate subject-level, parametrically-weighed “early”, “middle”, and “late” regressors for each pill condition; these were further contrasted to yield late>early maps that served as our primary measure of interest: the RE-related brain activity over the course of extinction, in line with standard practices^21,56^. We also averaged RE-related brain activity across all task runs to improve power and detect potential voxelwise correlations with next-day drug-biased behavior, and further explore potential associations with Table 1 cocaine use severity measures.

For fMRI analyses of RE-related activity over the course of extinction, subject-level maps were entered into group-level mixed-effects analyses using FSL’s FLAME 1& 2 (FMRIB’s Local Analysis of Mixed Effects), which improves variance estimations using Markov Chain Monte Carlo simulations for better population inferences^57^. All analyses were restricted to our *a priori* defined region of interest, the vmPFC, derived from the Harvard-Oxford Cortical Atlas, thresholded at 25% probability. To minimize Type I error, we selected an *a priori* cluster-defining threshold (Z>3.1), corrected to a cluster-extent threshold of p<.05, as recommended^58^, and further applied a cluster size threshold (k=5) to the surviving clusters.

In psychophysiological interaction (PPI) analyses, we examined the functional connectivity between RE-related brain activity over the course of extinction and the amygdala. We concatenated the reminded and non-reminded task events into a single timecourse and weighed the former as 1 and the latter as -1 to create the “psychological” regressor. We extracted reminded>non-reminded timeseries information using our vmPFC seed from each participant’s MPH- and placebo-related functional maps, yielding a “physiological” regressor. The interaction of these terms yielded a “PPI” regressor in our design matrix, to which we added all other regressors from the standard GLM described above to circumvent circularity and rule out non-task-specific coupling such as those due to anatomical connectivity^59^. Subject- and group-level statistics and cluster inferences for the PPI followed identical practices to those described above, with the exception that here, the results were restricted to a combined bilateral mask of the amygdala using the Harvard-Oxford Subcortical Atlas, thresholded at 25% probability. To further explore potential vmPFC functional connectivity targets that may be pertinent for salient drug cue extinction, we extracted connectivity strength information from the ventral tegmental area and the nucleus accumbens. The former was centered on coordinates (x=-3, y=-14, z=-19) that were previously associated with BOLD responses to salient drug cues in cocaine addiction^60^, and the latter was centered on coordinates (x=-14, y=3, z=-13) associated with reward and punishment anticipation^61^, tested via a 2 (pill) x 2 (phase: early, late) ANOVA for each region.

## Results

### Blood MPH Concentration Estimates

All participants yielded blood MPH concentrations that were congruent with the session where they received the MPH pill. Specifically, gas chromatography/mass spectrometry analyses indicated that samples acquired following the MPH pill administration yielded higher plasma MPH concentration (9.79 ug/L ± 4.28) compared to samples acquired following placebo (0.72 ug/L ± 0.42), *t*(15)=8.41, *p*<.001. See Supplementary Materials for details including quality control results.

### BOLD-fMRI Results

#### RE and Standard Extinction Effects under Placebo and MPH

Consistent with our expectation for a vmPFC impairment during standard extinction in iCUD^12^, we found a significant decrease (late<early trials) in non-reminded CS+ related vmPFC activity under placebo, t(17)=2.57, p=.020, Cohen’s d=0.60. Also consistent with our hypotheses, the reminder-only (reminded CS+ related activity under placebo) and MPH-only (i.e., non-reminded CS+ related activity under MPH) conditions both indicated modest effects: the former yielded a trend for an increase in vmPFC activity from early to late extinction, *t*(17)=1.70, *p*=.108, Cohen’s d=0.40 while the latter indicated no significant change, *t*(17)=0.18, *p*=.858, Cohen’s d=0.04; however, given the patterns in Figure 3, we explored this condition further via a one-sample t-test, and a significant change from implicit baseline was indeed evident when collapsed across all runs, *t*(53)=3.00, *p*=.002. Crucially, we found higher vmPFC activity during reminded vs. non-reminded CS+ related trials over the course of extinction under placebo as compared to MPH, Z=4.30, p=.004, 24 voxels, Cohen’s d=0.93, x=8, y=48, z=-12 (Figure 3). This interaction effect was driven by the combination of RE and MPH: a larger decrease in reminded CS+ related vmPFC activity from early to late extinction under MPH, *t*(17)=2.25, *p*=.038, Cohen’s d=0.53.

**Figure 3.**
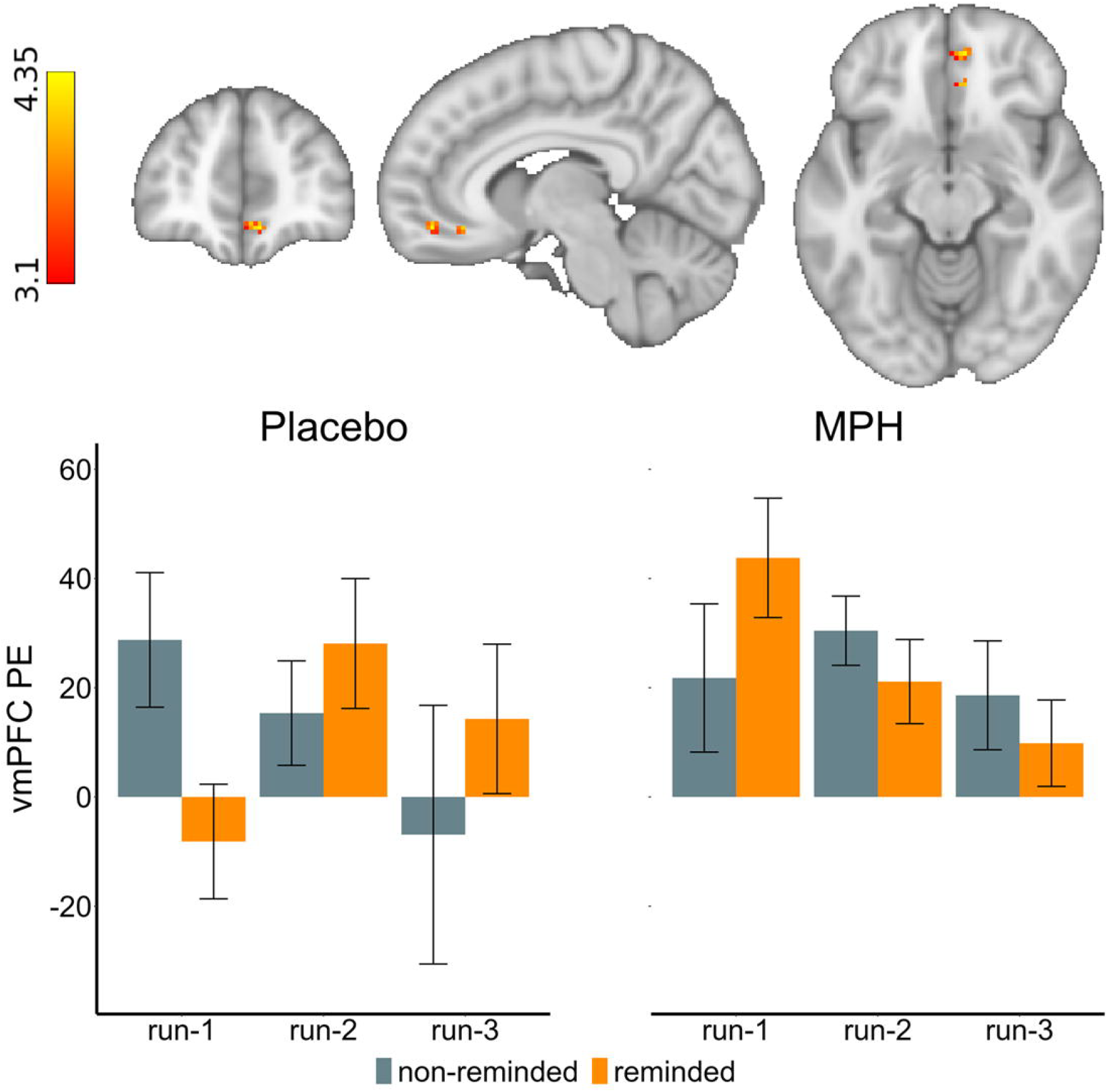
Individuals with cocaine use disorder (n=18) display impaired extinction-related ventromedial prefrontal cortex (vmPFC) activity under placebo, which normalizes under methylphenidate (MPH) enhanced retrieval-extinction (RE, i.e., reminded vs. non-reminded conditioned stimulus, or CS+ trials). We found higher vmPFC activity during RE over the course of extinction learning (i.e., run-3 vs. run-1) under placebo; a similar increase was noted for standard extinction (non-reminded CS+) during MPH. During MPH-enhanced RE vmPFC activity was decreased. The error bars reflect the standard error of the mean. Significant clusters are identified using a cluster-based Z-threshold of 3.1, corrected to p<.05, within an atlas-based mask of the *a priori* vmPFC region of interest. PE: parameter estimate.

#### Relationship between RE and Next-Day Drug-Biased Behaviors

The voxelwise correlations indicated a significant relationship only following the placebo pill, Z=4.06, *p*=.022, 8 voxels, Cohen’s q=0.68, x=8, y=48, z=-12, such that the higher the vmPFC signal in response to the reminded vs. non-reminded CS+, the more the drug vs. pleasant picture choices the next day (Figure 4). No significant clusters indicating brain-behavior correlations emerged under MPH.

**Figure 4.**
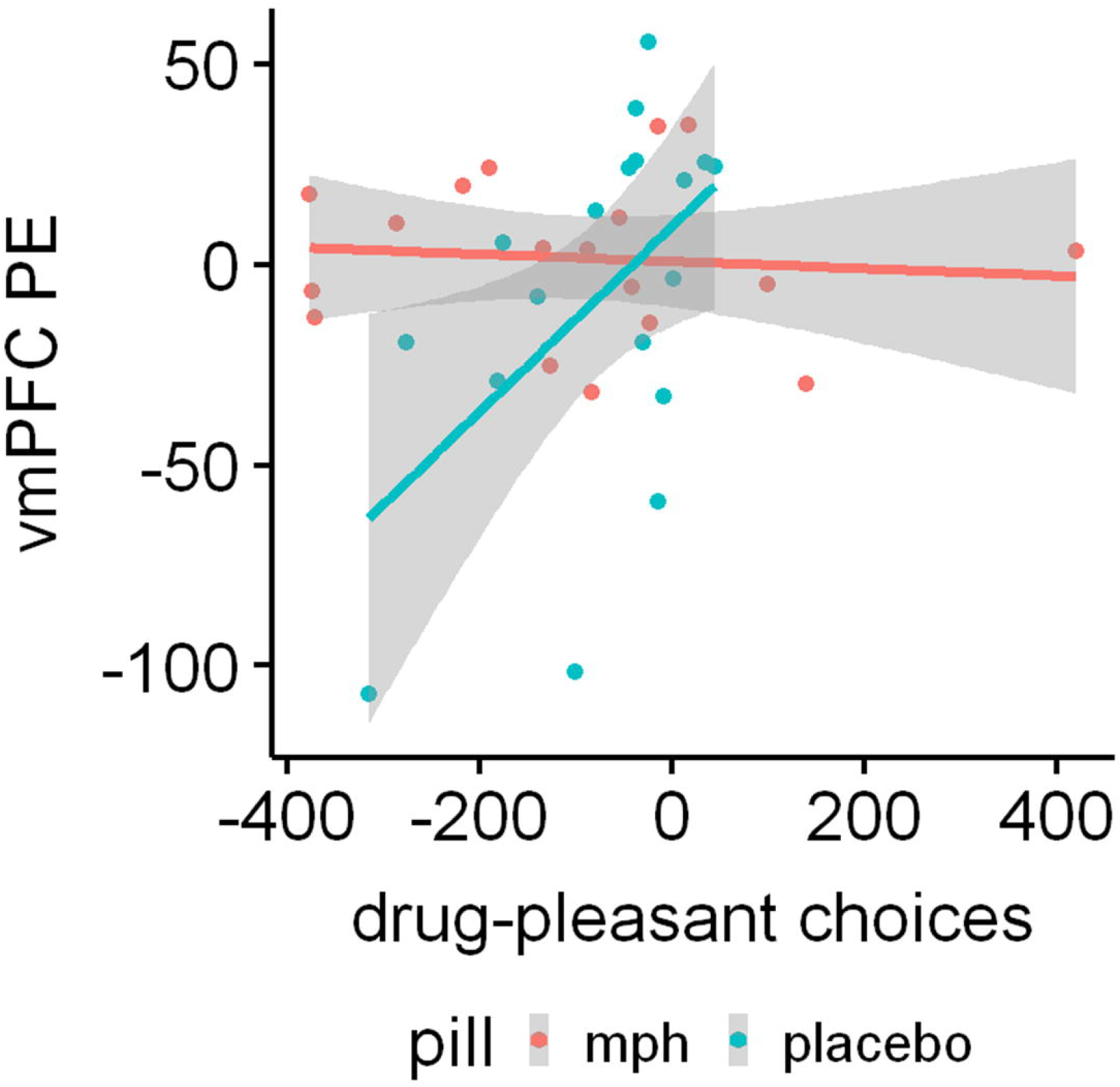
Higher mean retrieval-extinction ventromedial prefrontal cortex (vmPFC) activity under placebo correlates with more next-day drug-biased (compared to pleasant) choices in a picture-viewing task that estimates simulated drug seeking. This association is not evident under methylphenidate (MPH). Voxelwise correlation detected using cluster-based Z-threshold of 3.1, corrected to p<.05, within an atlas-based mask of the *a priori* vmPFC region of interest. PE: parameter estimate.

#### Functional Connectivity between the vmPFC and Amygdala

The PPI revealed a significant effect between the vmPFC and the left amygdala. Specifically, the strength of the functional connectivity during reminded vs. non-reminded CS+ between these regions showed a larger increase from early to late extinction phases under placebo compared to MPH, Z=4.07, *p*=.006, 21 voxels, Cohen’s d=0.836, x=-18, y=-4, z=-16 (Figure 5A). This effect was driven by a significant vmPFC-amygdala connectivity decrease from early to late extinction under MPH, *t*(17)=2.42, *p*=.027, Cohen’s d=0.57.

**Figure 5.**
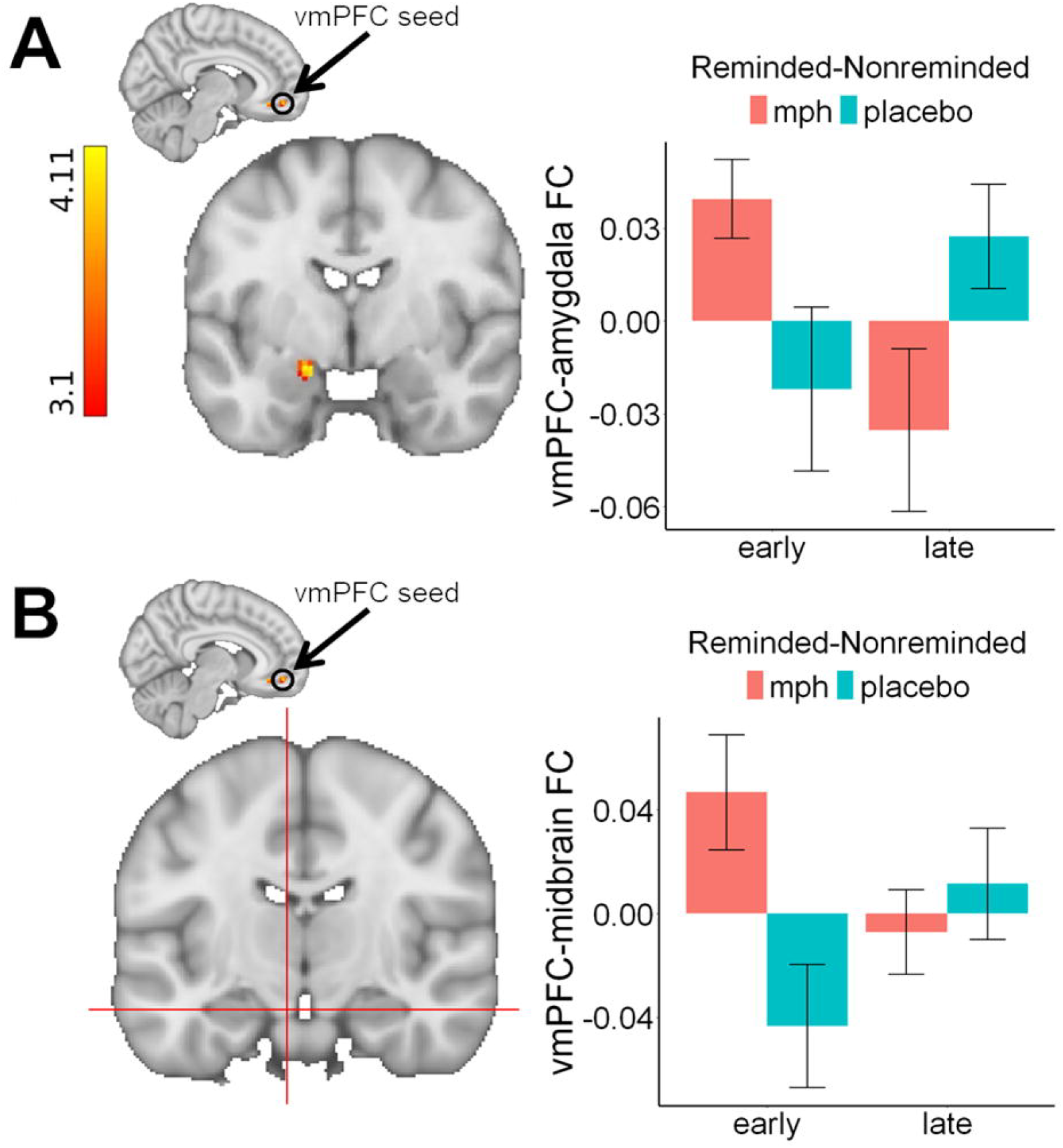
(A) A psychophysiological interaction illustrating differential functional connectivity between the ventromedial prefrontal cortex (vmPFC) seed and the amygdala. Functional connectivity (FC) increases during reminded vs. non-reminded CS+ between the vmPFC and amygdala over the course of extinction were higher under placebo compared to methylphenidate (MPH). Significant clusters are identified using a cluster-based Z-threshold of 3.1, corrected to p<.05, within an atlas-based mask of the *a priori amygdala* region of interest. (B) An exploratory psychophysiological interaction illustrating differential functional connectivity between the ventromedial prefrontal cortex (vmPFC) seed and the midbrain/ventral tegmental area during reminded compared to non-reminded conditioned stimulus (CS+) trials over the course of extinction.

#### Functional Connectivity between the vmPFC and Ventral Tegmental Area and Nucleus Accumbens

Exploratory analyses of the connectivity strength extracted from our primary PPI analyses indicated a significant functional connectivity effect between the vmPFC and ventral tegmental area during reminded vs. non-reminded cues over the course of extinction, *F*(1,51)=7.06, *p*=010, η2=0.12 (Figure 5B). Similarly to the amygdala, this effect was driven by trends of increases in connectivity under placebo, *t*(17)=1.94, *p*=.069, Cohen’s d=0.46, and decreases under MPH, t(17)=1.82, *p*=.086, Cohen’s d=0.43, from early to late extinction. The same analysis of the nucleus accumbens yielded no significant effect, *F*(1,51)=1.31, *p*=.258.

#### Exploratory Analyses of Relationships between RE and Drug Use Severity Measures

The voxelwise correlational explorations between reminded vs. non-reminded CS+ brain activity and the drug use severity measures indicated a significant relationship between the vmPFC activity under placebo vs. MPH and years of regular cocaine use, Z=3.76, *p*=.037, 7 voxels, x=2, y=38, z=-10, such that the higher the mean RE vmPFC activity under placebo, the more the years of regular cocaine use (Supplementary Figure 1). Under MPH, a significant relationship was evident with years of heaviest cocaine use, Z=3.71, *p*=.027, 7 voxels, x=2, y=48, z=-10, such that the lower the mean RE vmPFC activity under MPH, the more the years of heaviest use (Supplementary Figure 2). No other drug use severity correlations yielded significant clusters.

## Discussion

Our results revealed that neural deficits during standard extinction in iCUD can be modestly altered over the course of extinction by RE and single-dose MPH challenge separately. Crucially, it is when these behavioral and pharmacological interventions were combined that the effect was similar to that observed in the general population. Specifically, we revealed decreasing vmPFC activity in response the non-reminded CS+ under placebo (i.e., standard extinction). The reminded CS+ (i.e., RE) during placebo increased these vmPFC activations from early to late extinction phases (at a trend level), and MPH showed an overall higher activation levels to the non-reminded CS+ (using one-sample t-tests when comparing the collapsed extinction phases to the implicit baseline). Crucially, combining RE with MPH significantly reduced reliance on the vmPFC, such that its activity decreased over the course of extinction; vmPFC functional connectivity with the amygdala (and ventral tegmental area in exploratory analyses) was also normalized (decreased). The clinical relevance of these results was evident by a significant relationship between higher vmPFC RE activity only under placebo (but not MPH) with next-day drug-seeking. Further exploratory analyses indicated that higher vmPFC RE activity under placebo was also associated with more years of regular cocaine use while lower vmPFC RE activity under MPH was associated with more years of heaviest use. For the first time, these results document the neural substrates of a combined pharmacological-behavioral modulation of drug cue memory in human cocaine (or any) drug addiction.

The decreasing vmPFC recruitment as a function of standard extinction training (to the non-reminded cues under placebo) signifies an impairment in vmPFC-guided processes in drug addiction. In contrast to increases in vmPFC activity during standard extinction that are evident in the general population^5^, this result was expected based on our previous similar results obtained in a separate sample of iCUD using an entirely different extinction fMRI task (and imaging protocol)^12^. In interpreting this effect, we consider vmPFC’s role in value tracking. A large-scale meta-analysis of fMRI studies that assessed reward and punishment signals revealed that the vmPFC specifically encodes positive valuation (i.e., higher BOLD response to rewarding vs. punishing outcomes)^62^. This vmPFC role aligns with its increasing recruitment by fear extinction in the general population, thought to reflect the updating of the valuation of the CS+ as it dissociates from the negatively-valenced US. Given the similarly negatively-valenced drug US in our task, the decreasing engagement of the vmPFC over the course of standard extinction in iCUD is indicative of an impairment in tracking valuation. This analysis is further supported by the current CS valence ratings that were unchanged with extinction (i.e., not more positive). This impairment may also be reflective of ineffective exertion of control, consistent with preclinical evidence for cortical control over conditioned responses that originate in the amygdala^63^. Specifically, excitatory inputs from the infralimbic cortex (a vmPFC homologue) activate intercalated cells in the amygdala that inhibit output neurons in the central nucleus (the sub-region that drives the expression of conditioned responses)^64–66^, as also demonstrated by the reduction of conditioned responses via the electrical stimulation of the infralimbic cortex^67^.

Importantly, the use of reconsolidation alone (during placebo) or MPH alone (during standard extinction) modestly increased vmPFC responses, pointing to behavioral and pharmacological approaches that, in isolation, induce some (compensatory) overreliance on the vmPFC. Crucially, the combined effect of MPH and RE had a robust and normalizing effect on the vmPFC in iCUD. Specifically, during MPH and over the course of extinction, we revealed decreasing vmPFC activity during reminded trials, with parallel decreases in its limbic connectivity, suggesting disengagement of the vmPFC from the amygdala (and the dopaminergic ventral tegmental area) over the course of RE (where the cortical control may no longer be needed). These vmPFC activity and connectivity decreases with MPH-enhanced RE in the iCUD are similar to the (fear) reconsolidation effects previously documented in the general population, whereby the vmPFC is disengaged/bypassed during RE^5,21^. In line with this interpretation of normalization (decreases) of vmPFC function (and connectivity) with MPH-enhanced RE are our behavioral correlation results. Here, associations between increased vmPFC RE function and increased next-day drug seeking were evident only during placebo, suggesting a sustained conditioned response not observed during MPH. In exploratory analyses with drug use history a similar correlation emerged between increased vmPFC RE activity during placebo with longer regular cocaine use. In contrast, the more the decreases in vmPFC RE activity with MPH, the longer the heaviest use. These results suggest that individual differences in severity of use need to be taken into account for intervention development, where RE alone could be effective for those with longer cocaine use while MPH-enhanced RE could target those with extended periods of heavy use. These correlation results (and the separate effects of both conditions) await replication in larger samples.

Thus, we extend the limited clinical evidence for pharmacological supplementation of RE in drug addiction. The beta-adrenergic antagonist, Propranolol, was associated with reduced cue-induced cigarette craving when administered inside of the reconsolidation window in smokers^33^ with promising (yet tangential) results also in heroin addiction^32^. Taking into account the paucity of these studies and the negative results for an NMDA receptor antagonist^31^, underscore the promising and translational effects of MPH in our results. Our successful pharmacological modulation of the underlying neural signature of RE is consistent with MPH-mediated improvements in 24 hour delayed memory recall^68^, and persistence of retention for incidental (e.g., episodic) and formal (e.g., trained) memory when administered after acquisition in healthy individuals^69^. Preclinically, MPH reduced conditioned responses despite insufficient extinction training in rats, in a reversible pattern upon the blockade of D1-family (and beta-adrenergic) receptors^43^, consistent with its dopaminergic role in learning and memory mechanisms^70^.

Indeed, the mechanisms underlying these MPH-enhanced RE normalization effects may involve dopamine-related, learning-induced cortico-limbic plasticity^40^, invoking this neurotransmitter’s key role in inferring the causes of meaningful stimuli^71^ (e.g., by assigning a causal relationship to the preceding reminded CS-no US retrieval event and its subsequent extinction). This dopaminergic interpretation also agrees with evidence in humans suggesting that MPH increases dopamine concentration as a function of the cue/task salience and motivation (i.e., food cue-induced hunger ratings, increased interest and motivation to complete math operations)^72,73^. In this vein, the involvement of dopamine in the decoupling of the CS from the negatively-rated drug US in our study is further supported by the decreased vmPFC-midbrain connectivity with MPH over the course of RE, as remains to be verified using more direct estimates of dopamine neurotransmission.

Our results should be interpreted in light of some limitations. First, the inspection of potential sex- or treatment-related effects was not possible in this proof-of concept study, warranting larger-scale replications with balanced samples; it would be especially important to compare across drug classes (e.g., stimulants vs. opiates) and to study the potential impact on results of drug co-use. However, note that a cross-over design, as used here, reduces inter-subject variability, maximizing statistical power and providing a more precise neural comparison of learning and memory processes under a pharmacological challenge. Second, we did not include a healthy control group, as the neural signal during the modulation of drug memories is exclusively relevant to people with drug addiction. Third, the self-reported CS and US valence, arousal and craving (drug US only) ratings did not yield pill or learning phase effects (and only showed the expected lower valence and higher arousal of the drug US). While these patterns may indicate an impairment in value tracking in iCUD, they may also suggest that these subjective and explicit ratings are not sensitive to the Pavlovian processes induced by the task. Also, while we acquired physiological (e.g., blood pressure) results before MPH and placebo administration, these measurements were not repeated until the next day, limiting inferences about immediate drug-induced effects. Nonetheless, our previous efforts with physiological estimates closer in time to pill ingestion suggested similar null pill differences (with the exception of diastolic blood pressure, which did not survive multiple comparison correction here)^45^. Lastly, our longitudinal design testing MPH-enhanced RE inspected associations with next-day drug-biased behaviors. Future efforts can expand on this time window to study longer-term (and repeated dosage) effects of memory modulation on drug-seeking behaviors.

We provide the first neurobiological evidence for pharmacologically-enhancing drug memory modulation to normalize extinction-related neural processing in people with drug addiction. Overall, our results suggest that while modestly effective separately, the combination of RE and single dose of the partial dopamine agonist MPH can robustly normalize the overreliance on the compromised vmPFC and its mesolimbic connectivity in iCUD. The clinical utility of the MPH-enhanced RE driven bypassing of the vmPFC is further supported by a sustained relationship between vmPFC RE function and next-day drug seeking under placebo only. Promoting the “forgetting” of disruptive memories in drug addiction may reduce drug seeking via a parallel change in salience attribution and enhanced self-control—two hallmark neuropsychological substrates in drug addiction. Current results could allow direct comparisons with animal studies, promoting additional mechanistic approaches (e.g., molecular imaging or targeted brain stimulation) to alter or even erase maladaptive memories for enhancing treatment options in drug addiction. The role in these results of dopamine-related, learning-induced cortico-limbic plasticity, especially as related to inferring the causes of meaningful stimuli, remains to be explored. Thus, our findings contribute to developing the foundation for effective, synergistic neuropsychopharmacological strategies that ameliorate the persistent and cyclical pathology of drug addiction en route to enhancing recovery.

## Data Availability

All data produced in the present study are available upon reasonable request to the authors

## Acknowledgments

We thank Yui Ying Wong and Maggie Boros for their assistance in recruitment and data collection. This work was supported by grant no. T32DA053558 to A.O.C. as trainee, and grant no. R21DA054281 to R.Z.G. The funders had no role in the design and conduct of the study, the collection, management, analysis and interpretation of the data, the preparation, review or approval of the manuscript, and the decision to submit the manuscript for publication.

## Competing Interests

J.H.N.: consultant/advisory board for AGB Pharma, Cingulate Therapeutics, Hippo T&C, Lumos, Medice, Mentavi Health, MindTension, Otsuka, Signant Health and Supernus; research support from Cingulate, MindTension, and Supernus; honoraria for disease state lectures from Apsen and Otsuka, and served as a consultant for the US National Football League. The other authors declare no competing interests.

## Notes

### Clinical Trial

NCT05978167

### Author Declarations

The Icahn School of Medicine at Mount Sinai Institutional Review Board approved all study procedures. All participants provided written informed consent prior to participating in the study and were compensated after study completion.

