## Supplementary Materials for "The neural signature of methylphenidate-enhanced memory disruption in human drug addiction: a randomized clinical trial"

**Exclusion Criteria**

Exclusion criteria were as follows: 1) history of a cardiovascular (including high blood pressure and cardiac arrhythmias), neurodevelopmental (e.g., autism), neurological (including history of migraines or seizures), or a psychotic disorder, or psychiatric symptoms (e.g., panic attacks, anxiety disorder) that can be exacerbated by methylphenidate (MPH); 2) head trauma with loss of consciousness for more than 30 min; 3) any medication use (within 6 months) that may alter cerebral function (excluding some psychotropics common to individuals in treatment for drug use disorders) or that is known to interact with MPH (blood pressure medications, blood thinners, anti-epileptics, etc.); 4) current medical illness and/or evident infection; 5) abnormal ECG or vital signs at time of screening as determined by trained staff; 6) any other medical conditions that may alter cerebral function, specifically, endocrinological (including metabolic), oncological, or autoimmune diseases; 7) history of glaucoma; 8) being pregnant or nursing; 9) MRI contraindications; 10) reported hypersensitivity to MPH, motor tics, or family history or diagnosis of Tourette’s syndrome; and 11) being court-mandated to seek treatment for drug abuse and/or prisoners. For generalization purposes, we did not exclude participants with comorbidities that are highly prevalent in individuals with cocaine use disorder (iCUD) (but that are not specifically contraindicated to receiving MPH) (e.g., other drug or alcohol use disorders, depression, post-traumatic stress disorder).

**Neuroimaging Acquisition and Preprocessing**

Anatomical T1-weighted image acquisition parameters were as follows: 3D magnetization-prepared rapid gradient echo sequence (field of view: 256 × 256 × 179 mm^3^), 0.8 mm isotropic resolution, TR=2400 ms, TE=2.07 ms, TI: 1000 ms, flip angle: 8° with binomial (1, −1) fat saturation, bandwidth: 240 Hz/pixel, echo spacing: 7.6 ms, and an in-plane acceleration (generalized autocalibrating partially parallel acquisitions) factor of 2. The T1-weighted scan acquisition time was approximately 7 min. The blood-oxygen-level-dependent (BOLD) fMRI scans were acquired using a T2*-weighted, single-shot multiband accelerated (factor of 7) gradient-echo echo-planar image sequence: TE=35 ms, TR=1,000 ms, 2.1 mm isotropic resolution, 70 axial slices without gaps for whole-brain coverage (147 mm), field of view: 206 × 181 mm, matrix size: 96 × 84, flip angle: 60° (approximately Ernst angle), blipped Controlled Aliasing in Parallel Imaging Results in Higher Acceleration phase-encoding shift=field of view/3, bandwidth: 1860 kHz/Pixel with ramp sampling, echo spacing: 0.68 ms, and echo train length: 84 ms. The extinction task was administered over three, approximately 8 min functional runs. The 1 hour scan session included a resting-state scan after the extinction task that will be reported separately.

Raw BOLD-fMRI data were first converted to NIFTI and preprocessed using the fMRIprep pipeline version 20.2.3^1^ We applied intensity normalization and skull-stripping on to structural images using ANTS^2^. These images were then spatially normalized to the ICBM 152 Nonlinear Asymmetrical template via ANTS’ nonlinear registration^3,4^. We applied brain tissue segmentation via FSL’s FAST^5^ to estimate white matter, gray matter, and cerebrospinal fluid. Motion correction was carried out via FSL’s MCFLIRT, and susceptibility distortion correction was applied using spin-echo field maps that were acquired in opposing phase encoding directions via AFNI’s 3dQwarp^6,7^. These motion- and distortion-corrected images were co-registered to the participant’s structural images using boundary-based registration with 9 degrees of freedom via FSL’s FLIRT^4,8^. The correction, transformation, and registration steps were integrated into a single step workflow using ANTS, resampled to 2 mm isotropic resolution voxels. For each BOLD scan run, we extracted the following confounds: six translation and rotation parameters (x, y, and z for each) as motion regressors, global cerebrospinal fluid and white matter components, and cosine regressors for high-pass filtering (128 s cutoff) to ignore low-frequency drift related to scanner and physiological noise. The preprocessed data were spatially smoothed using a Gaussian kernel (5 mm full-width at half maximum, further smoothed to 10 mm full-width at half maximum for analyses that inspected cortical-to-subcortical connectivity as recommended^9^) to improve signal to noise ratio.

**Subjective US and CS Valence, Arousal, and Craving Ratings**

We performed 2 (Pill: MPH, Placebo) x 3 (Phase: Pre-conditioning, Post-conditioning, Re-exposure) x 2 (Cue: Drug, Neutral) linear mixed models with subject as a random factor for valence and arousal ratings, and a 2 (Pill) x 3 (Phase) linear mixed model for craving ratings with subject as random factor to determine participants’ subjective responses evoked by the US. We also performed a 2 (Pill) x 3 (Phase) x 3 (Stimulus: Reminded CS+, Non-reminded CS+, CS-) linear mixed model with subject as a random factor to inspect subjective valence and arousal responses elicited by the task CS.

Using US valence as the dependent variable (DV), we found a main effect of Cue [Neutral>Drug; *F*(1,174)=16.84, *p*<.001; degrees of freedom calculated using Pinheiro & Bates’ formula for mixed models^10^], but no main effects of Pill, Phase, or any interactions (all ps>.138), suggesting that the drug US was overall rated to be less pleasant than the neutral US, but did not change based on type of pill or learning phase. The US arousal DV yielded a main effect of Cue [Drug>Neutral; *F*(1,174)=51.67, *p*<.001], with no main effects of Pill, Phase, or their interactions (all ps>.186), suggesting that the drug US was rated to elicit higher arousal compared to the neutral US, but this pattern did not relate to type of pill or learning phase. The US craving DV indicated a non-significant trend for the main effect of Pill [Placebo>MPH; *F*(1,79)=3.86, *p*=.053], with no main effect of learning phase or interactions (all ps>.318), suggesting learning phase did not significantly affect drug US induced craving. The CS valence ratings as DV indicated a main effect of Stimulus [Non-reminded CS+>CS-; *F*(2,269)=4.25, *p*=.017], with no main effects for Pill or Phase, or their interactions (all ps>.108), supporting success of the conditioning. CS arousal ratings as DV yielded no main effects or interactions (all ps>.174). These patterns suggest that overall, the drug US was rated as more negatively valenced and arousing than the neutral US, that craving was slightly lower under MPH, and that the non-reminded CS+ cue was rated as more positive than the CS- (with the reminded CS+ being statistically comparable to both), with no other differential valence or arousal ratings based on the pill administered or the learning phase.

**US and CS Forced Choice Ratings**

We performed a 2 (Pill) x 3 (Phase) linear mixed model with subject as a random factor for forced choices between the drug and neutral US, using as DV averaged dummy coded choices made between these stimuli (0=neutral US, 1=drug US, such that larger numbers indicate more drug compared to neutral US choices). This analysis yielded no main effects of Pill or Phase, and no Pill x Phase interaction (all ps>.203), indicating that stimulus choices did not differ based on learning phase or type of pill administered. Targeting CS choices, for each combination of stimulus choice pairs (i.e., CS- vs Reminded CS+, CS- vs. Non-reminded CS+, Reminded CS+ vs. Non-reminded CS+, each dummy coded as 0 vs 1, respectively), we performed separate 2 (Pill) x 2 (Phase) linear mixed models with averaged choices as DV. These analyses yielded no main effects or interactions involving Pill or Phase for any of the stimulus comparisons (all ps>.169), indicating that choices between stimulus types did not differ based on pill or learning phase.

**Supplementary Table 1.** Affective, physiological, and drug use recency variables at both pill administration days.

|  | **Placebo** | | **Methylphenidate** | |  |
| --- | --- | --- | --- | --- | --- |
| **Variable** | **Mean** | **Std. Dev.**^1^ | **Mean** | **Std. Dev** | ***p*** ^2^ |
| Positive Affective State ^3^ | 24.33 | 11.21 | 27.78 | 10.3 | .22 |
| Negative Affective State ^3^ | 12.33 | 3.55 | 13.39 | 4.72 | .149 |
| Previous Night Sleep Hours | 6.58 | 1.71 | 6.16 | 2.44 | .504 |
| Previous Night Sleep Quality | 2.06 | 0.64 | 2 | 1.08 | .79 |
| Systolic Blood Pressure (pre) ^4^ | 121.33 | 13.24 | 121.5 | 13 | .967 |
| Systolic Blood Pressure (post) ^4^ | 121.89 | 14.6 | 124.24 | 15.57 | .344 |
| Diastolic Blood Pressure (pre) | 80.11 | 10.9 | 82.94 | 8.66 | .142 |
| Diastolic Blood Pressure (post) | 81.17 | 9.65 | 84.94 | 9.54 | .028 |
| Heart Rate (Beats/Minute) (pre) | 74.67 | 11.33 | 75.22 | 11.51 | .802 |
| Heart Rate (Beats/Minute) (post) | 79.72 | 12.64 | 80.29 | 13.08 | .646 |
| Pill Guess % Correct | 0.67 | 0.49 | 0.78 | 0.43 | .331 |
| Heroin Days Since Last Use ^5^ | 385.4 | 498.5 | 397.73 | 487.32 | .398 |
| Heroin Past 30 Day Use (Days) ^5^ | 0.07 | 0.26 | 0 | 0 | .334 |
| Cocaine Days Since Last Use ^6^ | 319.88 | 434.19 | 318.24 | 429.42 | .508 |
| Cocaine Past 30 Day Use (Days) ^6^ | 0.18 | 0.73 | 0 | 0 | .332 |
| Alcohol Days Since Last Use ^6^ | 1372.86 | 1911.57 | 1392.71 | 1961.18 | .383 |
| Cigarette Days Since Last Use | 0.12 | 0.34 | 0.71 | 2.66 | .386 |
| Cigarette Past 30 Day Use (Days) | 29.38 | 2.5 | 29.12 | 2.64 | .669 |

^1^ Std. Dev: standard deviation.

^2^ T-test comparisons between placebo and methylphenidate study days in each measure, familywise error corrected (α=.05/20=.002).

^3^ Acquired via the Positive and Negative Affective States questionnaire immediately after the MRI scan.

^4^ Pre and post measures acquired three hours before and the day after pill administration, respectively (during daily medical screening procedures). One participant’s post measures (in methylphenidate arm) not included due to missingness.

^5^ Data for heroin-related variables did not include two participants due to lack of heroin use history and one participant due to missingness.

^5^ Data for cocaine and alcohol-related variables did not include one participant due to missingness.

**Supplementary Table 2. Urine Toxicology**

| Substance | Placebo | Methylphenidate |
| --- | --- | --- |
| Amphetamine | 1/18 | 1/18 |
| Barbiturates | 0/18 | 0/18 |
| Benzodiazepines | 0/18 | 0/18 |
| Buprenorphine | 2/18 | 2/18 |
| Cannabis | 2/18 | 1/18 |
| Cocaine | 0/18 | 0/18 |
| Ethyl Glucuronide | 0/18 | 0/18 |
| Fentanyl | 3/18 | 1/18 |
| MDMA ^1^ | 0/18 | 0/18 |
| Methadone | 14/18 | 15/18 |
| Methamphetamine | 0/18 | 0/18 |
| None | 2/18 | 2/18 |
| Opiates ^2^ | 1/18 | 0/18 |
| Phencyclidine | 0/18 | 0/18 |

Complete toxicology reports from both pill administration days. Substances are categorized per drug test kit panel specifications (model: HCDOAV-6145EF1).

^1^ Methylenedioxymethamphetamine.

^2^ Other than fentanyl.

**Blood MPH Concentration Estimates: Methods and Quality Control Assessments**

A total of 32 samples from 16 participants were available for the analysis (two participants’ blood samples were insufficient in volume). Sample pairs were randomly assigned to analytical batches (n=4), accompanied by six quality control samples. A 100uL aliquot of the plasma sample was fortified with deuterated internal standard mix prior to extraction. MPH was extracted from samples using solid phase extraction (HLB prime, 3cc, Waters). Sample extracts were derivatized using N-methyl-N-(trimethylsilyl)trifluoroacetamide and N-Methyl-bis(trifluoramide) before analyzing them in gas chromatography/tandem mass spectrometry (Agielnt 7010B MS/8890 GC, Agilent Technologies). Three microliters of the extract was injected in the multimode inlet of the GC equipped with 30 m DB-XLB capillary column. Helium was used as the carrier gas at a constant flow of 1mL/min. The MS was operated in the election ionization (EI) mode at -70eV. Data acquisition was performed using a highly precise and accurate Multiple Reaction Monitoring method, and the isotope dilution internal standard calibration method was used to determine the MPH concentration in the sample. Monitored quality control parameters were all with acceptable range: Coefficient of Variation of repeated analysis: 3%, recovery of spiked quality control samples: >96%, Method Detection Limit (estimated as 3 x standard deviation of 8 quality control samples analyzed along with study samples): 0.7ug/L (Limit of Quantitation determined as 3 x the Method Detection Limit). We conducted MPH stability analyses and did not observe degradation over the sample storage period.

**Supplementary Figures**

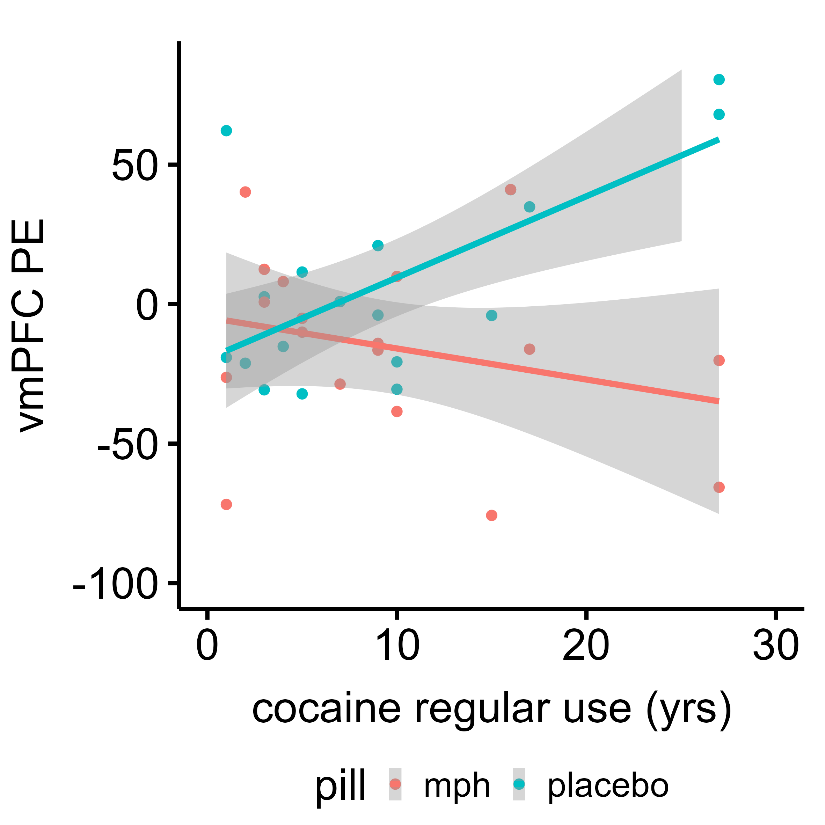

**Supplementary Figure 1.** Exploratory voxelwise correlation between retrieval-extinction ventromedial prefrontal cortex (vmPFC) activity and years of regular cocaine use. Higher mean retrieval-extinction vmPFC activity under placebo correlates with more years of regular cocaine use in individuals with cocaine use disorder. This association is not evident under methylphenidate (MPH). Voxelwise correlation detected using cluster-based Z-threshold of 3.1, corrected to p<.05, within an atlas-based mask of the *a priori* vmPFC region of interest. PE: parameter estimate.

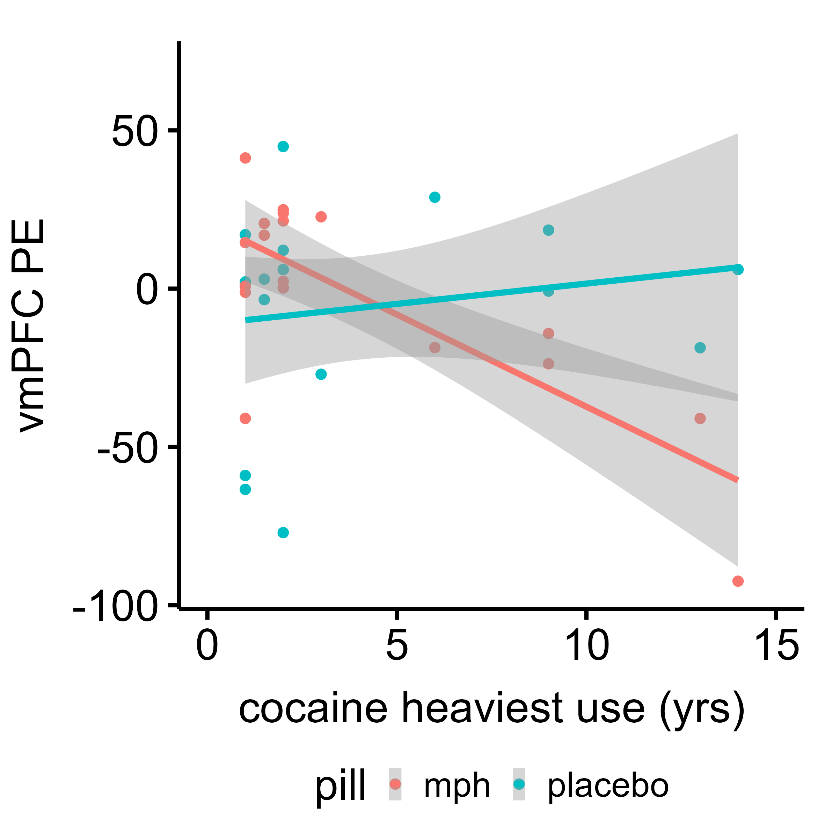

**Supplementary Figure 2.** Exploratory voxelwise correlation between retrieval-extinction ventromedial prefrontal cortex (vmPFC) activity and years of heaviest cocaine use. Lower mean retrieval-extinction vmPFC activity under methylphenidate (MPH) correlates with more years of heaviest cocaine use in individuals with cocaine use disorder. This association is not evident under placebo. Voxelwise correlation detected using cluster-based Z-threshold of 3.1, corrected to p<.05, within an atlas-based mask of the *a priori* vmPFC region of interest. PE: parameter estimate.
